# Project ECHO® for patients with chronic intestinal failure: Empowering people living with rare disease using a virtual telelearning model

**DOI:** 10.64898/2026.08.25.26361379

**Authors:** Kishore R. Iyer, Marion Winkler, Elisa Fisher, Vanessa Kumpf, Malvika Nair, Swapna R. Kakani, Kristy Poindexter, Andrew E. Jablonski, Emily Hoopes, Princess B. Ballog, Marjorie Nisenholtz, Rocco Friebel, Constantin T. Yiannoutsos, Joanne Lai, Kelly A. Tappenden

## Abstract

**Background:** Chronic intestinal failure is a devastating rare disease in which patients require complex and life-saving parenteral nutrition or intravenous fluids delivered through a central venous catheter. There is a shortage of clinical expertise to manage chronic intestinal failure and patients in the United States lack access to the limited number of expert care centers. We developed a patient intestinal failure (PIF) ECHO intervention with patient advocates who have lived experience with the goal of connecting patients and family caregivers virtually to multidisciplinary intestinal failure experts for best practice learning.

**Objective:** We pilot-tested the acceptability and feasibility of a direct-to-patient telelearning program based on the well-established ECHO® Model focused on best practices in chronic intestinal failure care.

**Setting and Participants:** 19 adults with chronic intestinal failure attended the pilot PIF-ECHO program for 12 consecutive weeks via Zoom between April and July 2026. All participants completed the post intervention questionnaire and 16 individuals participated in 3 focus groups. Design: A mixed methods evaluation was conducted. Questionnaires were assessed according to seven domains of the Theoretical Framework of Acceptability and qualitative data from the virtual focus groups were coded and analyzed using iterative thematic analysis. A data-derived PIF-ECHO logic model was developed to illustrate pathways between the program content and anticipated outcomes.

**Results:** There was strong or very strong agreement that sessions were accessible, enjoyable, worth the time spent, and improved understanding of intestinal failure and its management. Information learned increased confidence for self-advocacy in navigating healthcare needs, disease and therapy self-management, and improved well-being. Interaction with facilitators, expert presenters, and peers was positive, judgement free, validating, and respectful. Participants felt empowered and reported lower levels of emotional strain due to the supportive resources and knowledge gained.

**Conclusions:** A patient-facing tele-learning program in chronic intestinal failure is feasible, accessible, and acceptable to patients and appears to result in important short-term and medium- term benefits. The program was perceived as valuable and notably different from patient and peer-led support groups. The model could be applied more widely to other rare diseases.

**Lived Experience and Patient Contributions:** Four patient advocates with lived experience in chronic intestinal failure were involved throughout the study including pre-study interviews and focus groups to inform PIF-ECHO design and content, recruitment, as presenters on topics of self-advocacy and role of patient support groups, and in the analysis and refinement of the program logic model. Their input shaped the relevance and acceptability of the PIF-ECHO pilot program. All four patient advocates fulfil uniform requirements for authorship and are co-authors on this paper. This work documents a meaningful partnership in the creation of a patient-facing virtual tele-learning adaptation of the ECHO® model and establishes a valuable collaboration for future study of PIF-ECHO on a larger scale.

## Introduction

Chronic intestinal failure (CIF) is a devastating rare disease resulting from a reduction of gut function below the minimum necessary for the absorption of macronutrients and/or water and electrolytes, such that intravenous supplementation is required to maintain health and/or growth (1, 2). Patients with CIF require parenteral support (PS) in the form of parenteral nutrition (PN) or intravenous (IV) fluids delivered through an indwelling central venous catheter. While home PN (HPN) is life-saving for patients with CIF, HPN is often accompanied by life-threatening complications in the form of catheter related blood stream infection (CRBSI), or a syndrome of progressive intestinal failure associated liver disease (IFALD), apart from complications related to diarrhea, dehydration, fluid and electrolyte abnormalities which may lead to renal dysfunction (3-6).

The best outcomes for patients with CIF are achieved by teams of experts in multi-disciplinary teams (MDT) organized into intestinal rehabilitation programs (IRPs) (7-10). However, adult IRPs are few and far between and more than half the states do not have adult IRPs (11). One consequence of this is that patients frequently have to travel long distances to see their HPN provider to receive CIF care (12). With an estimated 25,000 adult patients receiving PN prescribed by just over 11,000 prescribers, there are significant disparities in access to care (12). We developed and administered a validated knowledge test in CIF to 100 US gastroenterologists (13). The results showed a concerning lack of knowledge of CIF care and HPN with no differences between self-described experts and non-experts (<u>13)</u>. To overcome the challenges of lack of access to expert care, we co-opted the well-established ECHO® Model in 2017 to launch the Learn Intestinal Failure Tele-ECHO program (LIFT-ECHO: https://liftecho.org/web/) to democratize knowledge and best practice learning in CIF (14-16). ECHO clinics use widely available video-conferencing technology to share clinical best practices, using case-based learning in an all-teach, all-learn environment. Since its inception we have conducted over 100 LIFT-ECHO clinics with an average of over 100 participating sites at each clinic from over 40 states and from 6 continents (16). We have observed limited engagement from non-expert community gastroenterologists and primary care physicians (PCP) at our LIFT-

ECHO clinics. Unlike widely prevalent diseases like Hepatitis C or Diabetes where an individual community physician might see a large number of affected patients, the average community physician may care for only one or two patients with a rare disease like CIF, in their entire career. Thus, they may not recognize the value, nor have the time to attend an educational program like LIFT-ECHO.

LIFT-ECHO, like every other ECHO program to date, excludes patients from participation in what is deemed a HIPAA-compliant platform. However, in rare diseases like CIF, patients often are more knowledgeable about their condition than many non-specialists. They are also the one common denominator across the many distinct and uncoordinated domains of US healthcare that they may traverse that range from specialists to PCPs or the local emergency room. We hypothesized that provision of multi-disciplinary education and support, directly to CIF patients through a live, virtual learning model – the Patient Intestinal Failure-ECHO Project (PIF-ECHO) – would enhance knowledge of best practices, increase confidence in self-care and capacity for self-advocacy and ultimately improve patient outcomes.

To test the feasibility, acceptability and perceived value of applying PIF-ECHO to address the lack of expertise in CIF nationally, we undertook formative IRB-approved qualitative studies to understand patient perspectives on the burden of CIF and on the value of adapting the ECHO Model to a patient facing context in CIF, supported by a pilot grant from AHRQ [1R03HS030321- 01; PI: Iyer, K]. (17). Four main themes emerged: 1) lack of provider access and support for patients living with CIF, 2) high levels of patient responsibility for disease management, self-advocacy, and system navigation, 3) limited access to patient-facing information on best practice CIF care, and, 4) severe emotional strain. Perceived value of PIF-ECHO centered on the program’s potential to improve knowledge and access to resources, which participants hypothesized would lead to improved capacity for disease self-management, and enhanced ability to advocate for their needs across healthcare settings. Findings informed the development of a pre-intervention PIF-ECHO logic model illustrating connections between program activities and anticipated outcomes (17). Based on the findings of the formative qualitative studies, a Pilot PIF-ECHO intervention was co-created with input from 4 patient advocates who brought lived experience to CIF.

## Materials and Methods

### Participant Recruitment

Consenting study participants meeting criteria for CIF on HPN were recruited into the study with help from patient advocates through the support groups administered by them on Facebook. Patients received modest incentives for their participation. The protocol was approved by BRANY IRB with letter number STUDY-24-01292 dated November 12, 2025. and by the New York Academy of Medicine Institutional Review Board (IRB). with the letter number #011426 dated January 20, 2026.

### Pilot PIF-ECHO Program Description

The Patient Intestinal Failure (PIF) ECHO was an IRP-approved pilot program comprising 12 consecutive weekly Zoom sessions (Table 1), 90 minutes in duration, designed to enhance best practices and confidence in self-care via a direct-to-patient adaptation of the ECHO® tele- learning model. The first 10 minutes included an overview of the program goals and speaker introductions, followed by a 20-25 minute expert presentation (including 1-2 pre and post test questions), 50-60 minute discussion with questions/answers, and 5 minutes of closing remarks. Participants were advised to use the chat function for contextual questions and to share their case-based experiences though the facilitators would invite them to unmute to present their questions and perspectives during the discussion session. The principal investigator (KI) and facilitators (MW, VK) welcomed the participants, reminded everyone that the goal of the research was to pilot-test a live, virtual learning system to connect patients and family caregivers with a team of intestinal failure experts and peers, and to determine how acceptable and valuable the sessions were. All research sessions and materials were confidential; de-identified recordings of the didactics were made available to review via a password protected portal. Participants were asked to be respectful of each other when speaking or sharing information and to maintain privacy. They were asked not to share names of physicians, hospitals, IRPs, or home infusion companies. Each week the facilitators reiterated that the speakers would not provide medical advice but would answer educational questions that would benefit everyone in the context of best practices. Speakers included gastroenterologists, surgeons, registered dietitian nutritionists, pharmacists, and clinical nurse specialists all of whom had expertise in chronic intestinal failure and home parenteral nutrition. One session included an overview of 4 patient support and advocacy groups presented by the group leaders or directors. The facilitators held a preliminary meeting with each speaker in advance of their session as an introduction to the pilot program and goals, and to review presentation slides and up to 3 key messages.

**Table 1.**
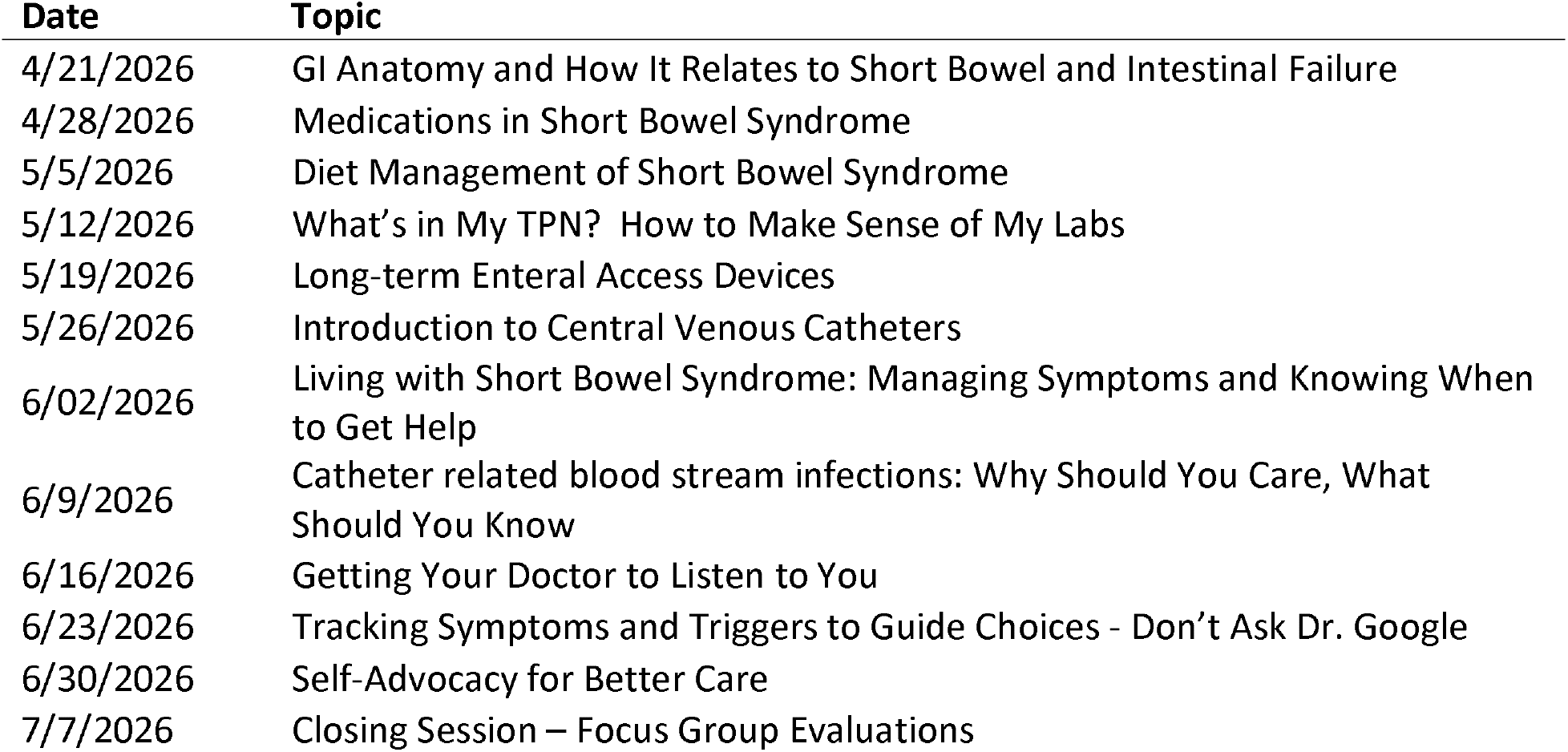
PIF-ECHO course content (Twelve weekly sessions)

### PIF-ECHO Program Evaluation

The evaluation of the pilot program was undertaken by a team of two researchers consisting of a lead evaluator with more than a decade of experience in mixed methods program evaluation and applied qualitative research methods (EF), and a supporting evaluator also trained in program evaluation and qualitative research (MN). It was supported by the PIF-ECHO implementation team, which included the PI leading the development and implementation of the PIF-ECHO program (KI) as well as a clinician with expertise in PN and quality of life (MW). This study used mixed methods, including a post-intervention survey and qualitative descriptive research (18) to: 1) assess feasibility and acceptability of the PIF-ECHO intervention; 2) explore patient perceptions of implementation of the PIF-ECHO intervention and perceived outcomes of participation; 3) examine consistency between participant perspectives and the program’s existing logic model, and 4) use findings to adapt the existing logic model to accurately reflect participant perspectives on the causal pathways connecting program activities to health outcomes, resulting in a robust theoretical framework that can be used to guide program implementation, improvement, expansion, and replication.

### Engagement of individuals with lived experience

The PIF-ECHO program has been co-created with individuals who have brought their lived experience to both the design and evaluation of the intervention. Four patient advocates with lived experience have been engaged in the design of the intervention. Focus groups with program participants were conducted prior to implementation and findings were used to inform program design, structure, and session topics. An initial draft of the logic model was shared with PIF-ECHO participants during a follow-up meeting; patient advocates supported interpretation of qualitative data and provided substantive feedback that was incorporated into the final logic model. All four patient advocates are co-authors meeting uniform requirements for authorship.

## Data collection

Participants were informed that participation in each of the evaluation activities described below was voluntary and they could end their participation at any time. Interested PIF_ECHO participants provided consent for participating in evaluation activities and received modest compensation.

### Survey

Evaluation and PIF-ECHO implementation teams collaborated to develop an online survey to assess program accessibility and acceptability. Accessibility questions focused on how and from where participants joined sessions. Acceptability was assessed according to seven domains identified in Sekhon et al.’s Theoretical Framework of Acceptability, 2017 (TFA), including affective attitude, burden, perceived effectiveness, ethicality, intervention coherence, opportunity costs and self-efficacy (19, 20). Surveys were administered via QuestionPro, an online survey platform. Responses were collected between June 30, 2026 and July 7, 2026. Average completion time was 6 minutes. All individuals who completed at least 1 PIF-ECHO session were invited to complete the survey. All PIF-ECHO participants completed the survey (n=19).

### Focus groups

The semi-structured focus group topic guide was developed collaboratively by the evaluation and PIF-ECHO implementation teams and contained questions exploring participant perceptions of accessibility and quality of the PIF-ECHO program, as well as perceived impact and recommendations for program improvement. All individuals who participated in at least one PIF-ECHO session were invited to join the focus group (n=19). Sixteen individuals participated in the focus groups, resulting in two groups with five participants and one group with six. The three groups were conducted on July 7, 2026 via Zoom teleconferencing breakout rooms. Focus groups were held during the regular PIF-ECHO meeting time via the same link that had been used for sessions to facilitate ease of access and participation. All groups lasted an average of 71 (range: 68-75) minutes.

All focus groups were led by a qualitative researcher trained in focus group facilitation who was primarily responsible for guiding the overall discussion. A co-facilitator responsible for notetaking, recording attendance, timekeeping, and addressing any technology-related issues also joined each group. To encourage honest and candid feedback, no one from the PIF-ECHO implementation team was present in focus groups; all focus group facilitators and co-facilitators were employed by the external evaluator.

## Data analysis

### Surveys

Survey data was downloaded from QuestionPro and transferred to STATA SE Version 17 for cleaning management, and analysis. Descriptive statistics were generated for all variables and reviewed by evaluation and PIF-ECHO implementation team members to examine responses across the domains of the TFA.

### Focus groups

Focus groups were recorded and professionally transcribed. Transcripts were managed and analyzed using Nvivo Version 15 software for qualitative analysis. Consistent with Braun and Clarke’s 2006 framework for reflexive thematic analysis and the qualitative descriptive approach described by Colorafi and Evans (18) data were analyzed using an iterative thematic analysis approach that involved multiple phases of review and interpretation by both evaluation and implementation team members.

Transcripts were initially reviewed by the evaluation team. The evaluation team then developed a codebook that included both pre-identified key topics, including those reflective of the program logic model developed prior to implementation described previously, as well as new topics that arose during focus group discussions or were identified during initial transcript reviews. Codes were then applied to all transcripts; coding was reviewed by both members of the evaluation team and multiple analytic discussions facilitated coding consistency. The evaluation and research teams initially developed a logic model for PIF-ECHO using data collected from patient advocates and patients prior to implementation on perceived need for and value of a patient-facing ECHO program for CIF (17). The evaluation team (EF, MN) shared preliminary findings with content experts on the PIF-ECHO implementation team (KI, MW) to facilitate clarification and refinement of the program logic model. This logic model was updated to reflect data on participant perspectives on activities, short-term, and intermediate outcomes collected for this study. The adjusted model was then used to create an updated coding scheme. Previously coded transcripts were updated using the refined coding scheme, allowing for further analysis of consistency between participant-reported short-term and intermediate outcomes and the updated logic model. By examining overlap between codes, this second round of analysis also facilitated identification of data extracts that support the causal pathways connecting program activities to outcomes illustrated in the model. The updated logic model as well as de-identified data extracts representative of short-term and intermediate outcomes were then shared back with program participants and four patient advocates with CIF lived experience to ensure that the model and interpretation of the data was aligned with participant perspectives. Feedback was incorporated into the final model. Figure 1 provides a visual representation of the PIF-ECHO program’s logic model, which illustrates hypothesized connections between program activities and outcomes.

**Figure 1.**
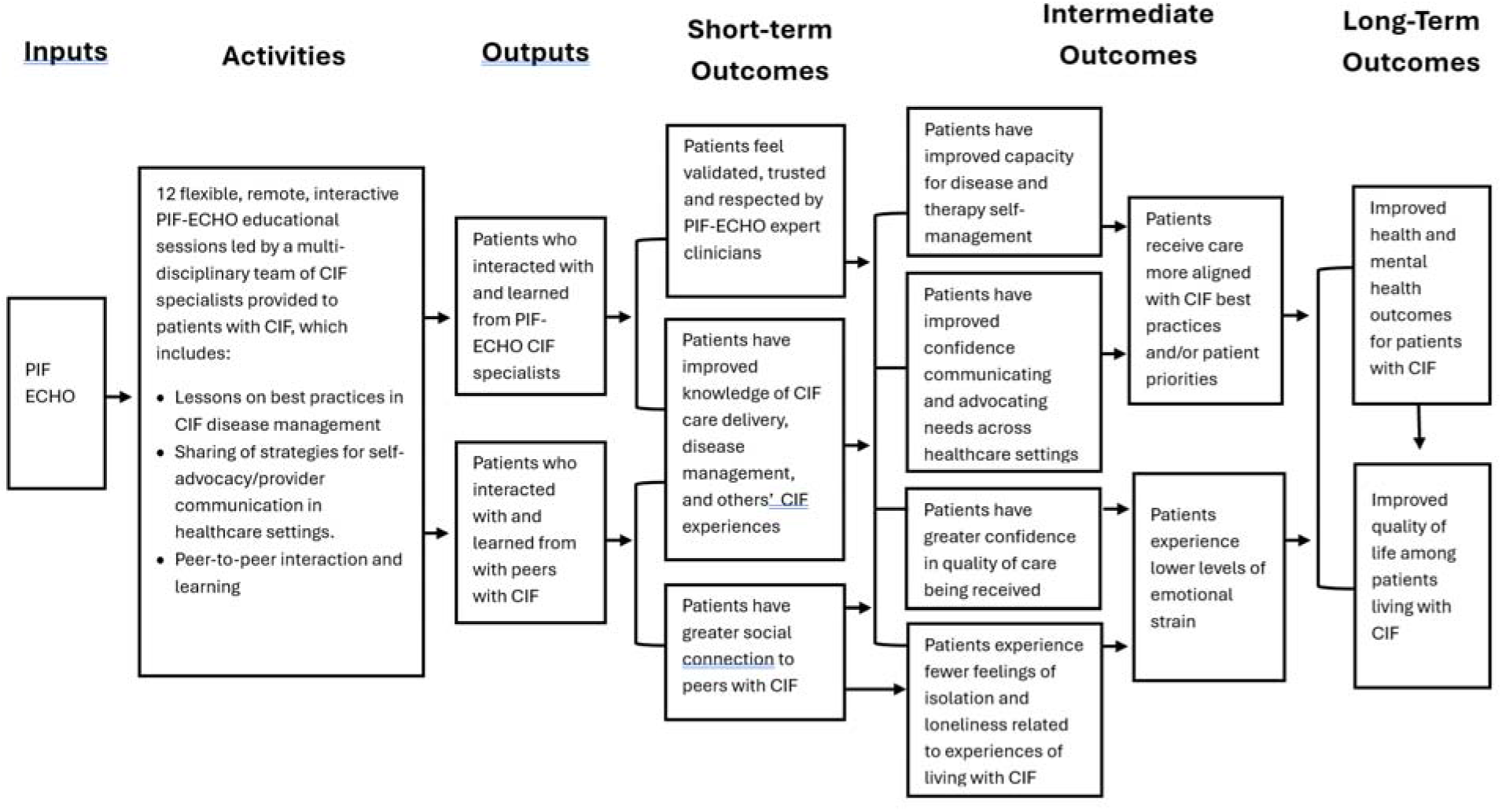
Patient-supported, data-driven PIF-ECHO logic model illustrating pathways between program activities and outcomes

Sample quotations pertaining to each theme and the pathways illustrated in the PIF-ECHO logic model are presented in the Results section. An ellipsis (…) is used to indicate omitted words or phrases, and brackets [ ] are used to designate words added for clarity.

## Results

### Participant Demographics

Nineteen adults signed informed consent to participate in the PIF-ECHO pilot program. The majority (n=16, 84%) identified as female and white (n=18, 95%), with a mean age of 47.3 ±13.8 years (23-77). All spoke English as their primary language. All participants self-identified as having CIF, though underlying conditions causing CIF varied. Participants reported many other medical co-morbidities; most commonly connective tissue/rheumatologic disease (n=9) and liver disease (n=7). While 9 patients (47%) reported having no hospitalizations in the last year, 10 reported being hospitalized for CIF-related complications (e.g., for central line infection, dehydration, diarrhea, vomiting); 7 (37%) had intestinal surgery in the last 2 years. All participants received home parenteral nutrition with mean length of therapy of 9.5 ± 11.8 years (<1 – 42) and the majority (n=15, 79%) infused additional intravenous fluids. A tunneled central line was the most (n=14, 74%) commonly reported venous access device. The most frequently reported insurance was Medicare (n=10), either as Medicare alone (n=4), Medicare and Medicaid (n=4), or Medicare and Private Insurance (n=2). Participants were restricted to and resided throughout the United States. Demographic characteristics for all PIF-ECHO participants are summarized in Table 2 and health characteristics pertaining to CIF and HPN are summarized in Table 3.

**Table 2.** PIF-ECHO participant demographics (N=19)

|  | **n** | **(%)** |
| --- | --- | --- |
| **Sex** |  |  |
| Female | 16 | (84%) |
| Male | 3 | (16%) |
| **Race/Ethnicity <sup>a</sup>** |  |  |
| White | 18 | (95%) |
| Hispanic or Latino | 1 | (5%) |
| **Age range** |  |  |
| 20 – 29 | 2 | (11%) |
| 30 – 39 | 3 | (16%) |
| 40 – 49 | 6 | (32%) |
| 50 – 59 | 5 | (26%) |
| 60 – 69 | 1 | (5%) |
| 70 – 79 | 2 | (11%) |
| **Highest grade or level of school completed** |  |  |
| Graduate degree | 6 | (32%) |
| Bachelor's degree | 5 | (26%) |
| Two-year degree (i.e., associate degree) | 4 | (21%) |
| Technical or vocational training | 2 | (11%) |
| Some college but no degree | 2 | (11%) |
| **Health insurance** |  |  |
| Private only | 6 | (32%) |
| Medicare only | 4 | (21%) |
| Medicare and Medicaid | 4 | (21%) |
| Medicaid only | 2 | (11%) |
| Private and Medicare | 2 | (11%) |
| Other | 1 | (5%) |
| **Geographic location** |  |  |
| West | 5 | (26%) |
| Northeast | 5 | (26%) |
| Midwest | 4 | (21%) |
| Southwest | 3 | (16%) |
| Southeast | 2 | (11%) |
| **Has caregiver** |  |  |
| No | 6 | (32%) |
| Yes | 13 | (68%) |
| **Relationship to caregiver <sup>a,b</sup>** |  |  |
| Spouse or partner | 9 | (69%) |
| Parent | 3 | (23%) |
| Adult child | 1 | (8%) |
| Another family member | 1 | (8%) |
| Paid professional (e.g., home health aide) | 1 | (8%) |

<sup>a</sup> Multiple responses permitted<sup>b</sup> Percentages based on number of respondents who indicated they had a caregiver (n=13)

**Table 3.** Health characteristics of PIF-ECHO participants (N=19)

|  | n | % |
| --- | --- | --- |
| **Primary diagnosis causing intestinal failure** |  |  |
| Short Bowel Syndrome (SBS) | 9 | (47%) |
| Intestinal Dysmotility | 3 | (16%) |
| Small Bowel Mucosal Disease and Intestinal Dysmotility | 2 | (11%) |
| Small Bowel Mucosal Disease and SBS | 1 | (5%) |
| Small Bowel Mucosal Disease and Crohn's Disease | 1 | (5%) |
| Intestinal Fistula and Intestinal Dysmotility | 1 | (5%) |
| Pseudo-obstruction | 1 | (5%) |
| Hirschsprung's Disease | 1 | (5%) |
| **Years on Home Parenteral Nutrition (PN)** |  |  |
| Less than 2 years | 4 | (21%) |
| 2 – 5 years | 5 | (26%) |
| 5 – 10 years | 4 | (21%) |
| 10 – 20 years | 3 | (16%) |
| More than 20 years | 2 | (11%) |
| Missing | 1 | (5%) |
| **Central venous access device** |  |  |
| Tunneled Catheter | 14 | (74%) |
| Peripherally Inserted Central Catheter | 4 | (21%) |
| Implanted Port | 1 | (5%) |
| **Receives care at Intestinal Rehabilitation Program (IRP)** |  |  |
| No | 14 | (74%) |
| Yes | 5 | (26%) |

### Activities and Outputs: Acceptability, Accessibility and Perceived Quality of the PIF-ECHO Program

#### Acceptability

Survey data were analyzed across the seven domains of the TFA to evaluate acceptability of the PIF-ECHO program. All participants agreed or strongly agreed that participation in PIF-ECHO sessions was enjoyable (affective attitude). The majority (n=16, 84%) agreed or strongly agreed that it was easy to find time to join PIF-ECHO, and nearly all (n=18, 95%) agreed or strongly agreed the time spent was worth it (burden). All agreed or strongly agreed that PIF-ECHO helped them to better understand their intestinal failure, nearly all (n=18. 95%) agreed or strongly agreed their understanding of the signs and symptoms of central line infections improved, and the majority (n=15, 79%) agreed or strongly agreed their understanding of how to care for their central line improved (perceived effectiveness). There was agreement or strong agreement (n=18, 95%) that it was important to have access to supportive resources for intestinal failure separate from their current healthcare team (ethicality). All participants agreed or strongly agreed that it was easy to understand how PIF-ECHO can help them personally (intervention coherence). The majority (n=14, 74%) changed their schedule to participate in PIF-ECHO; however most (n=17, 89%) disagreed or strongly disagreed that PIF- ECHO participation prevented them from doing other things they would prefer to be doing; a similar proportion (n=16, 84%) disagreed or strongly disagreed that their participation in PIF- ECHO caused them to spend less time with family and friends (opportunity costs). Nearly all (n=18, 95%) agreed or strongly agreed they had plans to use the information they learned in PIF-ECHO when caring for their central line, the majority (n=15, 79%) agreed or strongly agreed that PIF-ECHO increased their confidence for self-advocating in healthcare settings, all agreed or strongly agreed that PIF-ECHO helped them feel less alone in navigating their healthcare needs, and nearly all (n=18, 94%) agreed or strongly agreed they were confident that the information gained would improve their wellbeing (self-efficacy).

#### Accessibility

Seven of the 19 participants attended all 12 sessions and 17 of the participants attended 9 or more sessions. Most joined sessions from a laptop computer (68%), smartphone (53%), or a tablet (21%). All participants joined sessions from their home, though one-third (32%) also joined from a car, and one-quarter (26%) joined from a hospital/healthcare facility, and two participants (11%) joined from work. All survey participants agreed or strongly agreed that the technology to join the PIF-ECHO session was easy to use.

Overall, participants found the PIF-ECHO program to be highly accessible. They valued the virtual model as it allowed patients to participate regardless of geographic location and emphasized the importance of having the ability to join from any location convenient for them, including a hospital bed, car, or workplace. Barriers to participation included conflicting commitments (e.g., work, doctor appointments), caregiving responsibilities, time zone differences, and unanticipated health challenges. However, most participants explained that they attended PIF-ECHO sessions whenever possible.

> Sometimes people are in the hospital during these sessions, and they were participating. People are joining from all over the country in different types of situations. (Focus group participant)

#### Perceived Quality

Focus group participants discussed their interaction with PIF-ECHO facilitators, presenters and peers, and consistently shared positive perceptions of their experience in the PIF-ECHO program. They frequently highlighted the quality of facilitation, which was perceived as judgement-free and respectful, as well as the quality of the didactic presentations and information shared via presentations and during the question-and-answer portion of the sessions. Interacting with and learning from peers was also discussed.

> *I think [the facilitator] managed [sessions] as well as could be expected with all of us having so many different nuances to our health. So, I really appreciated how she tried to get to different people and group things together to address as many of the topics as possible. (Focus group participant)*

Nearly all survey participants (n=18, 95%) agreed or strongly agreed that the topics covered in PIF-ECHO were important to them as a patient. All survey participants reported that they would recommend PIF-ECHO to other individuals living with intestinal failure.

##### I. Participant Perspectives on Short-Term Outcomes of PIF-ECHO

Data collected from program participants provides strong support for short-term outcomes identified in the PIF-ECHO logic model. According to focus group participants, PIF-ECHO created a space where patients felt validated, trusted, and respected by participating CIF specialists. It also was perceived to result in gains in patient knowledge on CIF care delivery, disease management, and common patient experiences, as well as greater social connection to peers with CIF.

##### Ia. Feeling validated, trusted and respected by PIF-ECHO CIF specialists

Participants described experiences of feeling validated and respected during interactions with expert clinicians participating in PIF-ECHO, including both facilitators and presenters. They emphasized the value of—and for some, the novelty of—feeling heard and treated as trusted partners by healthcare providers with such deep knowledge and expertise in CIF. Some participants contrasted these experiences with those they have had with other healthcare providers who either made them feel disrespected, were too busy to answer their questions, or were less well-informed about their condition.

> *I’m very thankful to the people, all the people that gave presentations that we were treated as human beings, not as, “Oh, you’re just a patient and you’re nobody, you don’t know anything.” (Focus group participant)*

> *Because a lot of times we have doctors that won’t even address things. They don’t even want to hear it. So, I really felt listened to by the doctors [in PIF-ECHO]. (Focus group participant)*

##### Ib. Improved knowledge on CIF care delivery, disease management, and other patients’ CIF experiences

Participants consistently reported gains in knowledge as a result of their participation in PIF- ECHO, including information relevant to their clinical care as well as disease management. They highlighted lessons on topics that they considered particularly useful, learned new, or relevant to their case, and several participants reported taking notes or screenshots of the information presented. Those with less disease-related experience or more limited access to specialists frequently reported that information presented was brand new. Those with more experience or access to specialists reported learning some new or updated information.

> *I thought that the session on signs to look for infection was really helpful. I’m kind of new to all of this. I’m only about a year out on TPN, and I didn’t really get a lot of basic education, so I found that really helpful. (Focus group participant) Some of us were hearing that information for the first time even though we’ve been in treatment for years. Because we had the specialists. And I think having specialists was amazing; the doctors that came and spoke. (Focus group participant)*

While all participants felt they learned from the PIF-ECHO expert clinicians, many also emphasized the value of learning from their interactions with peers living with CIF. Participants explained that both groups provided access to expertise and experiences rarely available outside of PIF-ECHO. A few emphasized the difference between PIF-ECHO and support groups, attributed to the presence of expert clinicians and the psychological safety created by the program’s facilitators and structure.

> *During the discussion, you get to bring all that information together and see, “Okay, well, [this person is] different from me here, but she still has some similarities to me”…[Patients] benefit from the discussions by learning from other people like them. (Focus group participant)*

> *A lot of times, [patients participating in online support] groups give medical advice, and that’s not really sound. The person who runs it has nothing to do with it. It’s the [support group participants] who are just making the comments and stuff. So, I really enjoyed this because it was moderated and everything and so any things could be discussed. (Focus group participant)*

##### Ic. Greater social connection to peers with CIF

In addition to learning from other individuals living with CIF, participants frequently described the benefits of connecting socially during PIF-ECHO sessions. They valued the opportunity to develop relationships with others who could relate to the challenges inherent in managing CIF, noting that it is often difficult to meet others with shared experiences due to the rarity of the condition. Several described communicating or building friendships with other PIF-ECHO participants outside of sessions.

> *Just the camaraderie of seeing different people having to deal with their pumps and their lines during the session…But seeing people in those situations, everyone – we understand. (Focus group participant)*

> *I’ve made several connections with several people, and that was a bonus. I had a doctor’s appointment last week, so I couldn’t [come to PIF-ECHO]…so one of the friends I made here texted me, “You weren’t there. Are you okay?” (Focus group participant)*

##### I. Participant Perspectives on Intermediate Outcomes of PIF-ECHO

Focus group data also provides strong participant support for the intermediate outcomes described in the logic model. More specifically, participants connected short-term outcomes to improved capacity for disease and therapy self-management, improved ability to communicate and advocate for needs across healthcare settings; greater confidence in quality of care being received and reduced feelings of social isolation and loneliness related to CIF. They further linked these early intermediate outcomes to two additional intermediate outcomes: receipt of care that is more aligned with CIF best practices or patient priorities and reduced feelings of emotional strain.

##### IIa. Improved capacity for disease and therapy self-management

Improved self-efficacy related to management of their CIF was also commonly discussed by PIF- ECHO participants. They provided specific examples of how they expect to use - or have already used—information gained from PIF-ECHO to care for themselves, such as recognizing the signs and symptoms of infection, preventing or addressing dehydration, and managing diet. A few also noted that they shared information with a spouse or healthcare provider.

> *I kinda feel like more…empowered, and I feel a bit more comfortable knowing that I’ve been lucky so far that I haven’t had like infections, but I feel comfortable that if that does happen, I know what steps to take and how to bring up stuff to my doctors. (Focus group participant)*

> *Each week, getting to hear different topics and different people on their field of practice, I have learned a lot of things that I was able to share with my husband, things that I was able to kinda change in my own day-to-day. [For example, I am] making the oral rehydration drinks from scratch instead of relying on the products that are out there and kinda using them trial by error. (Focus group participant)*

##### IIb. Improved confidence communicating and advocating for needs across healthcare settings

Participants explained that information gained from trusted sources—including both CIF specialists and peers—helped them to feel more confident while communicating with healthcare providers and making personal healthcare decisions. They explained that being more knowledgeable and having their experiences validated by others with lived or professional experience helped them trust their instincts and advocate for their needs and priorities more confidently. In practice, this was important both when communicating with providers who were perceived as receptive to patient input but not well-informed, as well as with providers deemed less supportive.

> *[PIF-ECHO] gave me more confidence to know that I can stand my ground and say, “No. The reason I’m doing this well is because I’m on these treatments. I’m not willing to try to stop them right now. That’s not in my best interest or in my best interest for my quality of life.” And I didn’t have the confidence to feel really secure in saying that prior to these sessions, and I think that it helped me to have more confidence in that. (Focus group participant)*

> *I feel like it’s hard for me to speak up for myself. And I feel like this group was helpful in that and hearing other people [with CIF] and like, “No. You know your body, and this is right, and this is what I did.” I think that has been really helpful. (Focus group participant)*

##### IIc. Patients have greater confidence in quality of care being received

According to participants, learning information on CIF management from trusted experts and from the experiences of others helped them feel more confident that they were receiving the best possible or most appropriate care for their particular situation. Some explained that, while the information did not impact their care, they appreciated the confirmation and additional detail that learning from PIF-ECHO experts provided. Others reported that lessons learned from the expertise of providers in PIF-ECHO helped them to better manage their own care or make healthcare decisions that were more aligned with their own priorities.

> *I was gonna say, thinking back, I felt validated and reassured that the care I’m getting is good, especially after hearing what other people are experiencing. But, a lot of the recommendations that I had not formally been taught before are like, "Oh, my providers are doing that. Oh, they’re doing that." So, that was really reassuring for me. (Focus group participant)*

> *But if [the medication I am now taking due to information I learned from PIF-ECHO] works as they hope it will, it could really drastically change the quality of my life going forward. So, I’m just thankful to have [gotten] input from [a clinician] who has more experience than what I had locally. (Focus group participant)*

##### IId. Fewer feelings of isolation and loneliness related to experiences of living with CIF

Participants described the positive impact of forming social connections with other participants who could relate to their CIF struggles and hearing about experiences of others that were similar to their own, explaining that this camaraderie contributed to a reduction in the feelings of isolation and loneliness common among patients living with CIF. They also noted that interactions with CIF specialists through PIF-ECHO, who treated them with respect and took the time to provide education and answer questions, helped them to feel more supported and less alone while managing their complex condition.

> *Because you feel so isolated in the day-to-day…It’s like something is always missing when you’re with your friends that don’t get it…So, I always appreciate being around other people, being able to talk to them, and not having to explain all the little things. (Focus group participant)*

> *Getting some of that information [from a dietitian associated with my TPN provider] too, in addition to what I’ve learned with the PIF ECHO, has really made some favorable changes for me instead of just feeling like I’m on my own. (Focus group participant)*

##### IIe. Receipt of care aligned with CIF best practices and/or patient priorities

Participants discussed the various ways that participation in PIF-ECHO impacted their approach to managing their disease and working with their care team. By the end of the program, several participants explained that they had already used the knowledge or confidence they attributed to PIF-ECHO to make healthcare decisions or advocate for their own preferences with their care teams. Examples include trying a new medication, making decisions about surgery, and making changes to the way they manage their diet and hydration.

> *I think [PIF-ECHO] has been very helpful and just boosting my own confidence in feeling that I do know more about what’s going on and things like that… And I recently got a new GI team that is very against [my care preferences]. And so, I’m trying to hold my ground and protecting my stability, protecting my happiness, protecting the future that I see for myself. (Focus group participant)*

> *[PIF-ECHO] helped me make a new medication change decision that I’ve been grappling with for years now. So, I was grateful just for their willingness to answer questions. (Focus group participant)*

##### IIf. Lower levels of emotional strain

Participants—including those with extensive experience managing their illness and those with less—reported improvements in emotional wellbeing as a consequence of their participation in PIF-ECHO. They discussed feeling more calm, less depressed, and more hopeful about their future as a result of their participation in PIF-ECHO. Participants connected these changes to short-term and earlier intermediate outcomes, including feeling less alone and more empowered with both supportive resources and knowledge.

> *I’ve been sick for maybe 17 years, and it is the first time in 17 years that I felt hope and heard things from doctors that I could apply and I knew would make me feel better. (Focus group participant)*

> *A lot of times we’re gaslit. Our pain is dismissed, or our feelings, or our symptoms are dismissed, and having this right here where we have access to the education, I think it’s very important. It gives us, not just clarity, but mental and emotional peace. (Focus group participant)*

## Discussion

We report on a pilot feasibility study of a novel, direct-to-patient telelearning program based on the well-established ECHO® Model that has hitherto not been accessible to patients and/or caregivers. Our assessment using Sekhon et al.’s Theoretical Framework of Acceptability (19, 20) suggests that such an application of the ECHO Model in a patient-facing context is highly acceptable to patients; qualitative findings from PIF-ECHO participant focus groups indicate that patients found the PIF-ECHO pilot program to be accessible and high quality. Attendance in the PIF-ECHO sessions was consistently high with no patient drop-out, notwithstanding the many challenges patients face in their daily lives. In line with other ECHO® programs complete participation in all sessions was achieved by only 7 of 19 participants, though all 19 have continued in the ongoing extension phase.

Patients attributed multiple positive outcomes from PIF-ECHO. They reported improved knowledge on CIF care delivery disease management and in the medium term this led to improved capacity for disease and therapy self-management, one of the primary goals of the PIF-ECHO tele-learning program. Even within the limited 3-month horizon of this pilot study a small subset of patients reported being able to take ownership for major healthcare decisions from a position of empowerment and knowledge of best practices. Patients also note direct connections between program participation and fewer feelings of isolation and loneliness with improved peer connections and learning from others with similar but not identical lived experiences with CIF. This in turn led to lower levels of emotional strain.

One additional area merits further study. The beneficial outcomes accruing from the PIF-ECHO intervention were ascribed to facilitation of the program by clinical experts, didactic lectures and discussions with clinical experts, and experiences shared by peers living with CIF. Despite high perceived value of peer-to-peer learning, participants noted that these sessions functioned differently than patient/peer-led support groups. While the principle of “all teach, all learn” is intrinsic to the ECHO® Model, our evaluation was not designed to tease apart the relative contribution of the clinician-led teaching versus peer-to-peer learning. Insofar as ECHO programs are neither unduly prescriptive nor make any proprietary claims on the sources of learning within the model, the question may only be of academic significance.

Our application of the ECHO® Model in a patient-facing context in the rare disease, chronic intestinal failure and the resulting participant-endorsed program logic model may have much wider application to other rare diseases. There are rare instances of formal online tele-learning programs for patients and family care-givers (21, 22). To our knowledge, none has formally attempted to use the framework of the well-established ECHO® Model and there has not been any systematic evaluation of such programs.

Whether the short- and intermediate-term benefits seen from our limited Pilot PIF-ECHO program will translate to longer term, measurable improvements in clinical outcomes and quality of life remains to be proven; assessing causal impact will require a carefully randomized trial with a systematic implementation and evaluation approach. Such an approach will provide a more comprehensive understanding of core program components, program outcomes, and longer-term benefits and can help inform future efforts to address the problem of health inequities in rare disease stemming from a critical lack of expertise.

The value of patient and caregiver education in improving their care and ability to advocate is being increasingly recognized. Rare disease patients frequently carry the burden of self-education unaided, seeking online sources, social media and peer-support groups (23-25). A unique feature of the well-operationalized ECHO Model is that the model itself is agnostic to the domain and has been successfully applied not only to a range of chronic and widely prevalent diseases, but also to non-clinical areas such as education, climate change and emergency preparedness (26-30). Our preliminary report suggests that the ECHO® model can be safely and successfully adapted for patient-facing contexts in rare disease. If our preliminary results are validated in larger trials, we believe patient-facing ECHO programs in rare disease can help address the problem of health inequities stemming from a critical lack of expertise.

## Limitations

This study includes several limitations. First, recruitment for the PIF-ECHO program relied on self-selection among CIF patients already engaged in online social support platforms (i.e., Facebook groups), which may have introduced bias as volunteers 1) were already connected to an online community and 2) may have greater interest in virtual education and learning best practices than those in the broader population. Second, majority of the participants were white and female, which limits generalizability to other groups. While appropriate for a pilot program and feasibility study, reliance on qualitative data and the relatively small sample size further limits generalizability, as it cannot be assumed that the broader CIF population would share the perspectives of study participants. Thus, while findings are representative of the perspectives of the majority of PIF-ECHO participants to date, these encouraging results may not extend to individuals with CIF who are less engaged in their own care or are disconnected from online support resources.

## Conclusion

We report a qualitative impact evaluation of a pilot patient-facing tele-learning program in the rare disease, CIF, adapting the well-established ECHO® Model for a direct-to-patient context. Using the Theoretical Framework of Acceptability for pilot interventions, our study shows that such a patient-ECHO is feasible, accessible and acceptable to patients, is viewed as high-quality and appears to result in a range of short term and medium-term benefits. If these findings are proven in a larger scale randomized controlled trial, we believe the model could be applied more widely with minimal adaptation to other rare diseases.

## Data Availability

All data produced in the present study are available upon reasonable request to the authors

## Notes

### Competing Interest Statement

The authors have declared no competing interest.

### Clinical Trial

Not registered - limited pilot telelearning intervention

### Author Declarations

The protocol was approved by BRANY IRB with letter number STUDY-24-01292 dated November 12, 2025. and by the New York Academy of Medicine Institutional Review Board (IRB). with the letter number #011426 dated January 20, 2026.

